# OncoCTMiner: streamlining precision oncology trial matching via molecular profile analysis

**DOI:** 10.1101/2023.07.10.23292477

**Authors:** Quan Xu, Yueyue Liu, Dawei Sun, Xiaoqian Huang, Feihong Li, JinCheng Zhai, Yang Li, Qiming Zhou, Beifang Niu

## Abstract

**Summary:** OncoCTMiner is an innovative platform that streamlines precision oncology trial matching by integrating genetic profile analysis and clinical data. It utilizes manual tagging and automated entity recognition to identify six major biomedical concepts within clinical trial records. The platform currently contains a database of over 457,000 clinical trials, enabling quick and advanced search functionalities. Additionally, OncoCTMiner features an automated matching system based on genetic profiles and clinical data, providing real-time matching reports for suitable clinical trials. This platform aims to enhance patient enrollment in precision oncology trials, facilitating the development of personalized cancer therapies.

**Availability and Implementation:** OncoCTMiner is available at https://oncoctminer.chosenmedinfo.com.

**Supplementary information:** Supplementary data are available at *medRxiv* online.

**Graphic Abstract:** Graphic abstract:
A) OncoCTMiner’s role in precision oncology trial enrollment. B) OncoCTMiner takes clinical and genetic profiles as inputs and utilizes a trial matching and filtering system to generate a report of matched trials. C) Strategy for building the clinical trial eligibility criteria database. D) Automatic matching strategy for genomics-driven oncology trials.

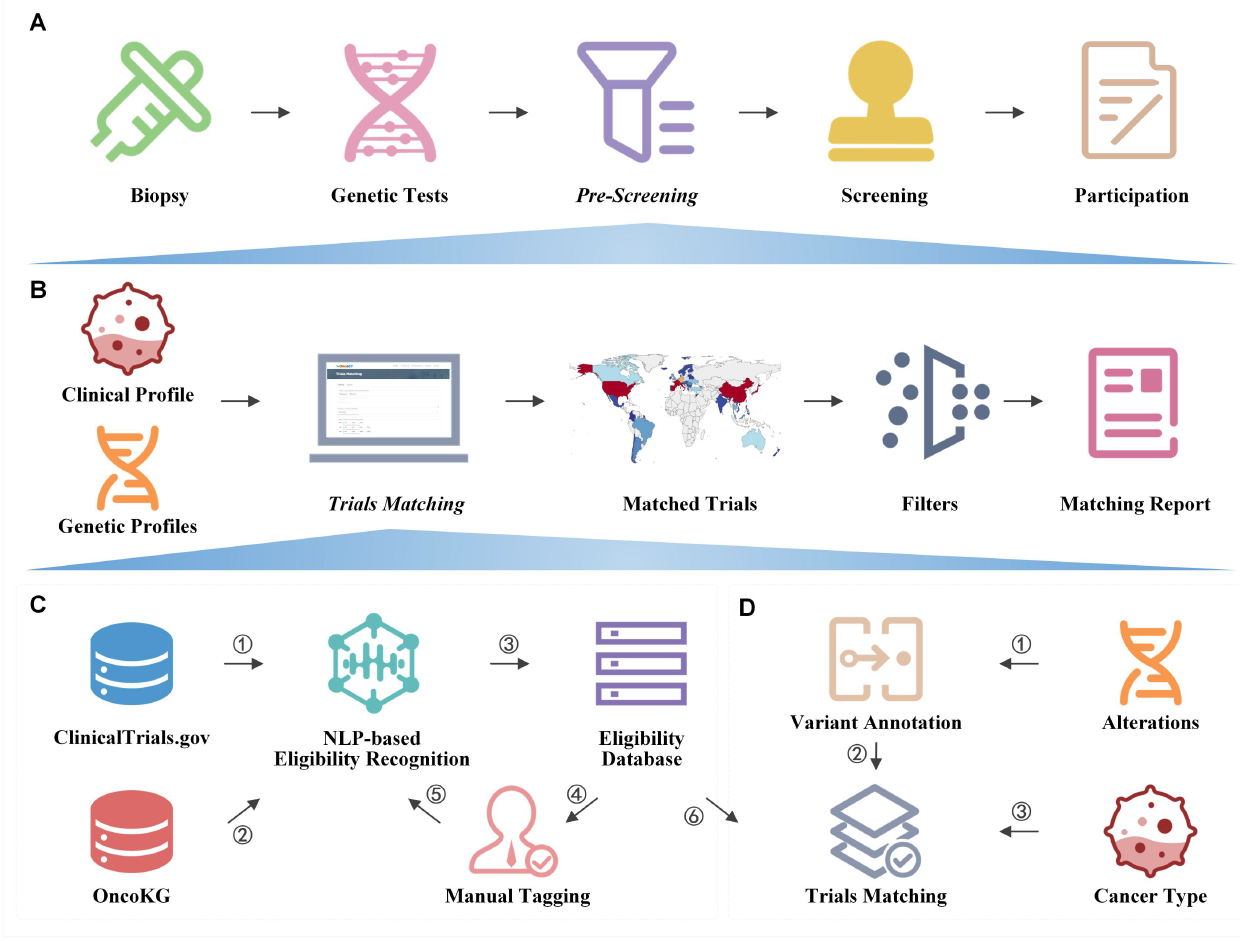

## Introduction

The integration of genetic profiling in cancer treatment has emerged as a promising avenue for targeted therapies and precision medicine-based approaches. However, realizing the full potential of these therapies necessitates active participation in clinical trials (Unger, et al., 2019). Unfortunately, the current participation rates in clinical trials, particularly in the domain of precision oncology, remain suboptimal (Jain, et al., 2021; Meric-Bernstam, et al., 2015; Stockley, et al., 2016). Several factors contribute to this issue, including limited physician knowledge, patient performance status, and concerns related to costs and attitudes (Ersek, et al., 2018). Furthermore, the process of connecting patient genetic data to trial eligibility criteria poses a significant challenge for physicians, who must navigate a multitude of trials to identify suitable options (Klein, et al., 2022).

To tackle these challenges, we present OncoCTMiner, an innovative platform specifically designed to streamline the process of precision oncology trial matching (Figure 1; Supplementary File 1). By leveraging the integration of genetic profile analysis and clinical data, OncoCTMiner offers real-time and automated capabilities for efficient trial matching. With a focus on user-friendliness, the platform aims to offer an intuitive solution that enhances automated trial matching. OncoCTMiner stands poised to make a substantial impact in the field of precision oncology, advancing research in personalized cancer treatments.

**Figure 1.**
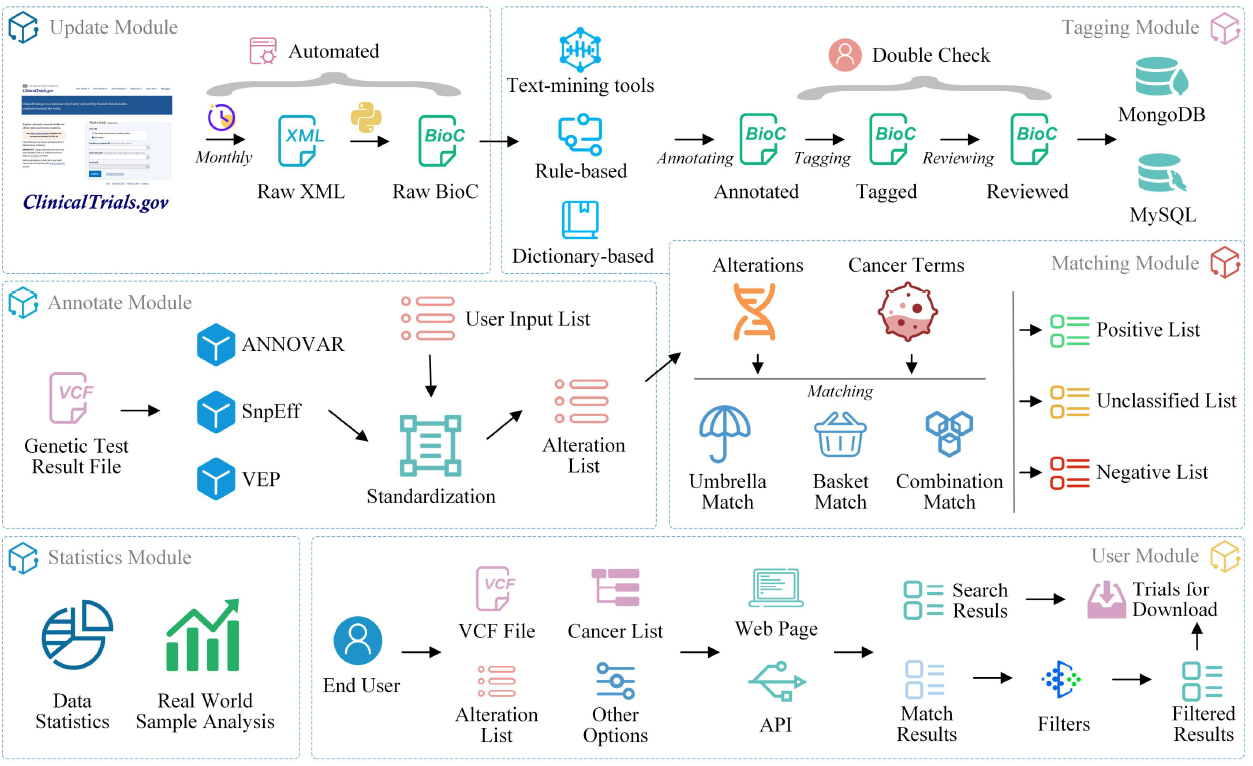
An overview of OncoCTMiner modules.

## Implementation

The implementation of OncoCTMiner involved several key steps to ensure its effectiveness and usability (Supplementary File 2, Text Mining). Firstly, clinical trial records were downloaded from ClinicalTrials.gov and converted to the BioC-JSON format for seamless interoperability and future data processing (Figure 1, Update Module). To enhance the accuracy and consistency of annotations, a platform was developed for manual tagging by biomedical experts (Xu, et al., 2022), incorporating a dual review mechanism and standard operating procedures (Figure 1, Update Module; Supplementary File 3). Open-source entity recognition tools (Leaman, et al., 2013; Leaman, et al., 2015; Wei, et al., 2015; Wei, et al., 2018) were leveraged and fine-tuned using the manually annotated corpora, enabling automated annotation of the clinical trials. All annotation results were aligned with standard terms for various entity types, such as genes, variations, diseases, and drugs, ensuring consistency and compatibility.

OncoCTMiner automates the matching of clinical trials based on the clinical and genetic profiles of tumor patients. It prompts users to provide clinical data and variant detection results, which are then annotated and matched against the eligibility criteria database. The system employs alteration annotation to map variations to standard alteration entries, increasing the positive matching rate and ensuring relevant trials are not missed (Figure 1, Annotate Module). Three matching modes, including basket, umbrella, and combination match, are utilized to categorize and filter the matched trials based on cancer types and alterations (Figure 1, Matching Module; Supplementary File 2, Trials Matching). Users can further filter the results based on specific criteria such as trial phase, recruiting status, and patient demographics. The matched and filtered trials are stored in MongoDB, and users can perform secondary filtering to obtain a satisfactory list of clinical trials meeting their requirements.

The OncoCTMiner system comprises a web application (APP) and multiple application programming interfaces (APIs). The web application was developed using SpringBoot, Mybatis-plus, LayUI, EasyWeb, and jQuery, providing a user-friendly interface. The APIs, built with the Flask-RESTful framework in Python, grant programmers access to clinical trial search functionalities in the BioC-JSON format. MySQL and MongoDB were employed for efficient database management.

## Results

The OncoCTMiner platform has yielded impressive results in mining oncology trial eligibility criteria and facilitating trial matching. The database currently encompasses a vast collection of 457,145 clinical trials, with 122,761 specifically related to cancer or containing cancer-related terms. Notably, 2,258 studies underwent manual double review to ensure data accuracy and quality.

These clinical trials were categorized into six major biomedical concepts, resulting in over 8.15 million entities and more than 9.32 million entity-criteria-trial triplets. OncoCTMiner provides intuitive and efficient search functions, allowing users to retrieve clinical trials based on keywords and biomedical concepts.

Moreover, the platform features an automated matching system that connects patients’ genetic profiles and clinical data to suitable clinical trials. The system receives variations called by the bioinformatics analysis pipelines as input for performing genetic profile-level trial matching. It also accepts clinical data for narrowing down clinical trial options. Upon analysis submission, OncoCTMiner generates matching reports which categorized trials as positive, negative, or unclassified. Positive trials indicate suitable options for consideration, negative trials are deemed unsuitable for participation, while unclassified trials require further evaluation.

OncoCTMiner has achieved remarkable results, showcasing its ability to rapidly retrieve information, improve precision oncology trial enrollment, and accelerate the development of potentially impactful tumor treatments. To enhance user comprehension, we provide a comprehensive web-based user guide (the ‘TUTORIAL’ page) that encompasses the platform’s main features. This includes step-by-step tutorials on clinical trial search and matching services, accompanied by relevant examples on the respective function pages.

## Conclusions

OncoCTMiner, a state-of-the-art platform, is specifically developed for mining oncology trial eligibility criteria, providing rapid and efficient information retrieval capabilities. Its sophisticated pre-screening functionality enables real-time matching of clinical trials based on patients’ genetic profiles and clinical data. The platform’s ability to enhance accrual rates for precision oncology clinical trials accelerates the development of potentially high-efficiency tumor treatments, ultimately benefiting a larger population of individuals affected by cancer.

## Data Availability

All data produced in the present study are available upon reasonable request to the authors

https://oncoctminer.chosenmedinfo.com/

## Funding

This work was supported by National Natural Science Foundation of China [grant number 92259101, 31771466], Strategic Priority Research Program of the Chinese Academy of Sciences, China [grant number XDB38040100], and the Cancer Genome Atlas of China (CGAC) project (YCZYPT [2018]06) from the National Human Genetic Resources Sharing Service Platform (2005DKA21300). The funders had no role in the design of the study, collection, analysis, interpretation of data, and in writing the manuscript.

## Notes

### Competing Interest Statement

The authors have declared no competing interest.

## References

Ersek, J.L., et al. Implementin. Precision Medicine Programs and Clinical Trials in the Community-Based Oncology Practice: Barriers and Best Practices. Am Soc Clin Oncol Educ Book 2018;38:188–196.

Jain, N.M., et al. Learnings From Precision Clinical Trial Matching for Oncology Patients Who Received NGS Testing. JCO Clin Cancer Inform 2021;5:231–238.

Klein, H., et al. MatchMiner: An open source platform for cancer precision medicine. medRxiv 2022:2022.2002.2002.22270186.

Leaman, R., Islamaj Dogan, R. and Lu, Z. DNorm: disease name normalization with pairwise learning to rank. Bioinformatics 2013;29(22):2909–2917.

Leaman, R., Wei, C.H. and Lu, Z. tmChem: a high performance approach for chemical named entity recognition and normalization. J Cheminform 2015;7(Suppl 1 Text mining for chemistry and the CHEMDNER track):S3.

Meric-Bernstam, F., et al. Feasibility of Large-Scale Genomic Testing to Facilitate Enrollment Onto Genomically Matched Clinical Trials. J Clin Oncol 2015;33(25):2753–2762.

Stockley, T.L., et al. Molecular profiling of advanced solid tumors and patient outcomes with genotype-matched clinical trials: the Princess Margaret IMPACT/COMPACT trial. Genome Med 2016;8(1):109.

Unger, J.M., et al. Systematic Review and Meta-Analysis of the Magnitude of Structural, Clinical, and Physician and Patient Barriers to Cancer Clinical Trial Participation. J Natl Cancer Inst 2019;111(3):245–255.

Wei, C.H., Kao, H.Y. and Lu, Z. GNormPlus: An Integrative Approach for Tagging Genes, Gene Families, and Protein Domains. Biomed Res Int 2015;2015:918710.

Wei, C.H., et al. tmVar 2.0: integrating genomic variant information from literature with dbSNP and ClinVar for precision medicine. Bioinformatics 2018;34(1):80–87.

Xu, Q., et al. OncoPubMiner: a platform for mining oncology publications. Briefings in Bioinformatics 2022.

