## Supplementary File 1 for "OncoCTMiner: streamlining precision oncology trial matching via molecular profile analysis"

**Supplementary Table S1: Comparison of OncoCTMiner to other systems.**

| Features \ Systems | OncoCTMiner | My Cancer Genome[1] | OCTANE[2] | MatchMiner[3] |
|---|---|---|---|---|
| *Terminologies / Ontologies* | | | | |
| Disease/Cancer | NCIt, MeSH, Disease Ontology, OncoTree | DiseaseOntology, NCIt, WHO, OncoTree, SNOMEDCT, UMLS | - | OncoTree |
| Gene | HGNC, NCBI-Gene | RefSeq, UTA | NCBI-Gene | - |
| Alteration | CIViC, CGI, PMKB, DoCM, OncoPDSS, ClinVar, COSMIC | HGVS, ISCN, UTA | - | - |
| Chemical/Drug | DrugCentral, ChEBI, ChEMBL, PharmGKB, DrugBank, | NCIt, NCIm, SNOMED CT, DrugBank, VANDF | NCIt | - |
| Biomarker | Self-built terminology | - | - | - |
| Therapy | Self-built terminology | - | - | - |
| *Data Related* | | | | |
| Trials Source(s) | ClinicalTrials.gov | NCI-Supported Clinical Trials, UMIN Clinical Trials Registry, ClinicalTrials.gov | ClinicalTrials.gov, CORe | ClinicalTrials.gov |
| Trails Content | Eligibility criteria, Meta data, Other detailed info | Eligibility criteria, Meta data, Other detailed info | Eligibility criteria, Meta data, Other detailed | Eligibility criteria, Meta data |
| *Entity-level criteria* | Yes | Yes | - | Yes |
| *Research Field* | Cancer | Cancer | Cancer | Cancer |
| *Clinical trials* | 436,009 | 9,809 | 5,439 | - |
| *Disease/Cancer* | | 955 | 252 | - |
| *Gene* | | 18,271 (gene + alteration) | 779 | - |
| *Alteration* | | | - | - |
| *Chemical/Drug* | | 2,861 | 1,453 | - |
| Trials Update | Monthly | Every two weeks | Daily | Daily |
| Data format | Structured, BioC-JSON based | Structured, webpage based | Structured，Oracle database | Structured, CTML based |
| *Functions* | | | | |
| Manual tagging function | √ | - | - | - |
| Trials searching | √ | √ | √ | √ |
| *Entity-based* | √ | √ | - | √ |
| *Metadata-based* | √ | √ | - | √ |
| Trials matching | √ | - | √ | √ |
| *Variant Annotation* | √ | - | - | - |
| *VCF-supported* | √ | - | - | - |
| *Patient-level* | √ | - | - | √ |
| Trials Filtering | √ | - | - | √ |
| Trials Download | √ | - | - | - |
| RESTful API | Yes, open and free | Yes, need license | - | - |
| Web-based open access | Yes | Yes | No | No |
| URL | https://oncoctminer.chosenmedinfo.com/ | https://www.mycancergenome.org/ | https://pct.mdanderson.org/octane/ | https://matchminer.org/ |

(to be continued)

(continue)

| DQueST[4] | Criteria2Query[5] | EliIE[6] | DCMS[7] |
|---|---|---|---|
| OMOP concepts | OMOP concepts | OMOP concepts, SNOMED-CT, LOINC | - |
| - | - | No | - |
| - | - | No | - |
| OMOP concepts | OMOP concepts | OMOP concepts | - |
| - | - | - | - |
| - | - | - | - |
| ClinicalTrials.gov | ClinicalTrials.gov criteria, user-entered criteria | ClinicalTrials.gov | MSKCC therapeutic clinical trials |
| Eligibility criteria | Eligibility criteria | Eligibility criteria | Eligibility criteria, Meta data, Other detailed info |
| - | - | - | - |
| Does not focus on specific field | Does not focus on specific field | Does not focus on specific field | Cancer |
| 252,330 | - | 230 (training set) | 840 |
| - | - | - | Not mentioned |
| Not mentioned | - | - | Not mentioned |
| Not mentioned | - | - | Not mentioned |
| - | - | - | Not mentioned |
| Not mentioned | - | - | Daily |
| Not mentioned | Structured, JSON based | Structured, XML based | Structured |
| - | - | - | - |
| √ | - | - | - |
| √ | - | - | - |
| Yes, dynamic questionnaire | - | - | - |
| No | - | - | √ |
| No | - | - | - |
| No | - | - | - |
| No | - | - | √ |
| √ | - | - | - |
| - | √ | - | - |
| - | Yes | - | - |
| Website unavailable | Website unavailable | - | No |
| https://impact.dbmi.columbia.edu/dquest-flask/ | http://www.ohdsi.org/web/criteria2query/ | - | - |

Note:
1 The comparative data of My Cancer Genome is derived from:
a) Holt, M.E., Mittendorf, K.F., LeNoue-Newton, M., Jain, N.M., Anderson, I., Lovly, C.M., Osterman, T., Micheel, C. and Levy, M. (2021) My Cancer Genome: Coevolution of Precision Oncology and a Molecular Oncology Knowledgebase. JCO Clin Cancer Inform, 5, 995-1004.
b) https://www.mycancergenome.org
2 The comparative data of OCTANE is derived from:
a) Zeng, J., Shufean, M.A., Khotskaya, Y., Yang, D., Kahle, M., Johnson, A., Holla, V., Sanchez, N., Mills Shaw, K.R., Bernstam, E.V. et al. (2019) OCTANE: Oncology Clinical Trial Annotation Engine. JCO Clin Cancer Inform, 3, 1-11.
b) https://pct.mdanderson.org
3 The comparative data of MatchMiner is derived from:
a) Klein, H., Mazor, T., Siegel, E., Trukhanov, P., Ovalle, A., Vecchio Fitz, C.D., Zwiesler, Z., Kumari, P., Van Der Veen, B., Marriott, E. et al. (2022) MatchMiner: an open-source platform for cancer precision medicine. NPJ Precis Oncol, 6, 69.
b) https://matchminer.org
4 The comparative data of DQueST is derived from:
a) Liu, C., Yuan, C., Butler, A.M., Carvajal, R.D., Li, Z.R., Ta, C.N. and Weng, C. (2019) DQueST: dynamic questionnaire for search of clinical trials. J Am Med Inform Assoc, 26, 1333-1343.
b) https://impact.dbmi.columbia.edu/dquest-flask
5 The comparative data of Criteria2Query is derived from:
a) Yuan, C., Ryan, P.B., Ta, C., Guo, Y., Li, Z., Hardin, J., Makadia, R., Jin, P., Shang, N., Kang, T. et al. (2019) Criteria2Query: a natural language interface to clinical databases for cohort definition. J Am Med Inform Assoc, 26, 294-305.
b) http://www.ohdsi.org/web/criteria2query
6 The comparative data of EliIE is derived from:
a) Kang, T., Zhang, S., Tang, Y., Hruby, G.W., Rusanov, A., Elhadad, N. and Weng, C. (2017) EliIE: An open-source information extraction system for clinical trial eligibility criteria. J Am Med Inform Assoc, 24, 1062-1071.
7 The comparative data of DCMS is derived from:
a) Eubank, M.H., Hyman, D.M., Kanakamedala, A.D., Gardos, S.M., Wills, J.M. and Stetson, P.D. (2016) Automated eligibility screening and monitoring for genotype-driven precision oncology trials. J Am Med Inform Assoc, 23, 777-781.

* The '-' in each cell indicates that the information is not availeble or that the feature does not exist.
