## Supplementary File 2 for "OncoCTMiner: streamlining precision oncology trial matching via molecular profile analysis"

**Text Mining**

*Data Loader*

ClinicalTrials.gov is a widely used database providing comprehensive information on clinical trials for both the general public and healthcare professionals (Zwierzyna, et al., 2018). To enhance interoperability and facilitate future data processing and exchange, we downloaded the ZIP file containing all study records in extensible markup language format from ClinicalTrials.gov and converted them to BioC-JSON format (Peng, et al., 2014) (Figure 1, Update Module).

*Manual Tagging*

We developed a platform for tagging clinical trials based on our previous work (Xu, et al., 2022) (Figure S1). Oncology trials involving gene, alteration, and drug entities are searched, screened, selected, and added to a list of pre-designed tagging projects, followed by double-checking by team members (Figure S1A- S1C). To ensure that individual annotators have a consistent reference standard and that identified bio-concepts are of high quality, we established a standard processing procedure (Supplementary File 3) for tagging entities.

OncoCTMiner aims to establish a comprehensive database of eligibility criteria for oncology trials and connect patients with suitable trials through a search engine and automated matching system. We use the 'minimization' principle for entity recognition to improve standardization, for example, dividing 'HER2-positive breast cancer' into an alteration and a cancer, and subdividing 'NSCLC with KEAP1, NFE2L2 and/or STK11 mutation' into a cancer type, three genes, and an alteration, which further normalized into 'KEAP1:mutation', 'NFE2L2:mutation' and 'STK11:mutation'.

OncoCTMiner distinguishes itself from comparable systems by not only tokenizing and normalizing biomedical concepts, but also determining whether an entity is a recruitment condition for a given clinical trial and its classification based on context. Eligibility criteria are classified into three types: 'not criteria' (NC), 'inclusion criteria' (inclusion) and 'exclusion criteria' (exclusion). Entities outside the eligibility criteria section are categorized as 'not available' (NA) since their context cannot be used to evaluate eligibility criteria. We apply the 'loose inclusion, tight exclusion' principle to minimize the false negative rate during trial pre-screening, while allowing for a relatively high false positive rate to facilitate further manual review of the pre-screened trial list.

*Entity recognition and standardization*

The system identified 6 categories of biological entities: disease/cancer, genes, alterations, chemicals/drugs, biomarkers and therapies. DNorm (Leaman, et al., 2013), GNormPlus (Wei, et al., 2015), tmVar2.0 (Wei, et al., 2018) and tmChem (Leaman, et al., 2015) were used to mine disease, genes, alterations, and chemical entities, respectively. Biomarkers are indicators discovered by genetic testing or immunohistochemistry that predict the efficacy of specific treatment regimens, such as TMB, MSI, and mismatch repair (MMR). Therapies refer to non-drug treatments, including drug treatment categories like 'chemotherapy' and 'immunotherapy'. We constructed two terminologies for recognition of biomarkers and therapies entities using a dictionary-based strategy.

Most entity annotation software matches recognized entities to commonly used databases. For instance, GNormPlus maps annotated genes/proteins to National Center for Biotechnology Information Gene identifiers, tmVar2.0 maps annotated variations to dbSNP RS identifiers, and DNorm and tmChem map annotated diseases/cancers and compounds/drugs, respectively, to Medical Subject Headings (MeSH) (https://www.ncbi.nlm.nih.gov/mesh/) identifiers. However, these standard identifiers do not cover all identified entities, requiring full standardization to facilitate later clinical trial retrieval and matching. We merged and built corresponding term sets for various entity types from several terminology and ontology databases, including OncoTree (Kundra, et al., 2021), DiseaseOntology (Schriml, et al., 2022), National Cancer Institute Thesaurus (https://ncit.nci.nih.gov/ncitbrowser/start.jsf), and MeSH. We gathered 55,558 cancer entries and established synonym connections or father-child relationships between each item, creating a unique cancer ontology. Using these terminologies or ontologies, we normalized all entities and mapped all synonymous terms to the same standard terms.

*Updates and archive*

Clinical trial enrollment status or enrollment criteria may be updated at any time. To ensure the system's accuracy, we update the trial database monthly, adding new clinical trials as they become available and updating existing trials as their content changes. This guarantees that users always have access to the most up-to-date clinical trial information. Historical versions of clinical trial data, particularly manually annotated data, will be archived as a corpus. As more data are gathered in the future, the entity recognition model will be fine-tuned and recognition efficiency will be constantly enhanced.

### Trials Matching

OncoCTMiner automates clinical trial matching based on the clinical and genetic profiles of tumor patients. Users are prompted to provide clinical data and variant detection results, which are then automatically annotated and matched against the eligibility criteria database. The variant annotation process involves identifying all detected variations in the user-provided variant call format (VCF) format data and mapping them to the standard entry of alterations. For trial matching, the system takes the cancer type selected by the user and the standard alteration terms as input and matches them against clinical trials in the eligibility database (Figure S2).

#### *Alteration Annotation*

The user-uploaded VCF file undergoes annotation by three software programs, VEP (McLaren, et al., 2016), ANNOVAR (Wang, et al., 2010) and SnpEff (Cingolani, et al., 2012), at the back-end of the system. The annotation results are then merged and mapped to the standard entries of alterations. Variation annotation not only matches specific mutations but also determines the type of variation that the mutation belongs to. For example, the mutation 'EGFR p.L858R' can match not only 'EGFR:L858R' but also 'EGFR:Activating mutations', 'EGFR:exon21mut', and 'EGFR:Mutations' (Xu, et al., 2020). This increases the positive matching rate of clinical trials standardized on these mutations in the system, reduces the chance of missing relevant trials, and offers more hope to patients.

#### *Trials matching and screening*

To match clinical trials, the system utilizes the cancer types and alteration list selected by the user as fundamental requirements. Three matching modes are provided: basket, umbrella, and combination match. Basket matching selects clinical trials that use variations as inclusion criteria (or NA) as the preferred conditions. These trials are then matched with cancer terms and categorized into negative, positive, and unclassified trial lists (Figure S2A). Umbrella matching is similar to basket matching, but with cancer and alteration listed as matching conditions in reverse order (Figure S2B). The goal of combination matching is to match both types of entries to the trials simultaneously. If either entity matches the trial, it will be instantly added to the provisional list for further categorization (Figure S2C). The list generated by these matching strategies is further filtered by user-specified conditions, such as clinical trial phase, recruiting status, trial center location, patient gender, and age, with only trials that meet the requirements being preserved (Figure S2D).

The clinical trials that are matched and filtered are stored in MongoDB in JSON format, utilizing the GridFS technology for space reduction due to the large amount of clinical trial data associated with plenty of matching jobs. Users can perform secondary filtering based on metadata and entity data, retaining trials that meet the criteria and removing those that do not. The final filtered list is saved in the same format, and can be re-screened by the user at any time to obtain a satisfactory list of clinical trials that meet their requirements in terms of both quality and quantity.

### REFERENCES

Cingolani, P., et al. A program for annotating and predicting the effects of single nucleotide polymorphisms, SnpEff: SNPs in the genome of Drosophila melanogaster strain w1118; iso-2; iso-3. Fly (Austin) 2012;6(2):80-92.

Kundra, R., et al. OncoTree: A Cancer Classification System for Precision Oncology. JCO Clin Cancer Inform 2021;5:221-230.

Leaman, R., Islamaj Dogan, R. and Lu, Z. DNorm: disease name normalization with pairwise learning to rank. Bioinformatics 2013;29(22):2909-2917.

Leaman, R., Wei, C.H. and Lu, Z. tmChem: a high performance approach for chemical named entity recognition and normalization. J Cheminform 2015;7(Suppl 1 Text mining for chemistry and the CHEMDNER track):S3.

McLaren, W., et al. The Ensembl Variant Effect Predictor. Genome Biol 2016;17(1):122.

Peng, Y., et al. iSimp in BioC standard format: enhancing the interoperability of a sentence simplification system. Database (Oxford) 2014;2014.

Schriml, L.M., et al. The Human Disease Ontology 2022 update. Nucleic Acids Res 2022;50(D1):D1255-D1261.

Wang, K., Li, M. and Hakonarson, H. ANNOVAR: functional annotation of genetic variants from high-throughput sequencing data. Nucleic Acids Res 2010;38(16):e164.

Wei, C.H., Kao, H.Y. and Lu, Z. GNormPlus: An Integrative Approach for Tagging Genes, Gene Families, and Protein Domains. Biomed Res Int 2015;2015:918710.

Wei, C.H., et al. tmVar 2.0: integrating genomic variant information from literature with dbSNP and ClinVar for precision medicine. Bioinformatics 2018;34(1):80-87.

Xu, Q., et al. OncoPubMiner: a platform for mining oncology publications. Briefings in Bioinformatics 2022.

Xu, Q., et al. OncoPDSS: an evidence-based clinical decision support system for oncology pharmacotherapy at the individual level. BMC Cancer 2020;20(1):740.

Zwierzyna, M., et al. Clinical trial design and dissemination: comprehensive analysis of clinicaltrials.gov and PubMed data since 2005. BMJ 2018;361:k2130.

### FIGURES

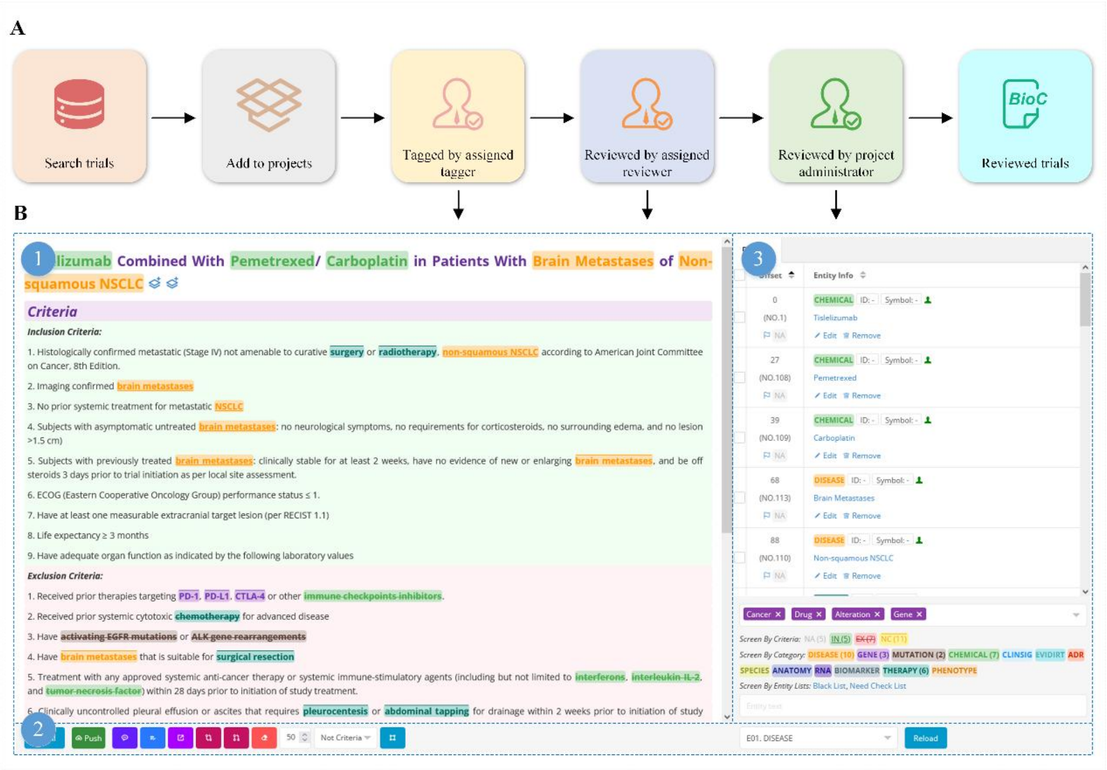


**Figure S1. Manual tagging of clinical trials.** A) Workflow for manual tagging of clinical trials; B) Overview of the trial tagging page: 1) Trial details with highlighted bio-concepts, 2) the manual tagging tools bar, and 3) real-time presentation of entity information.

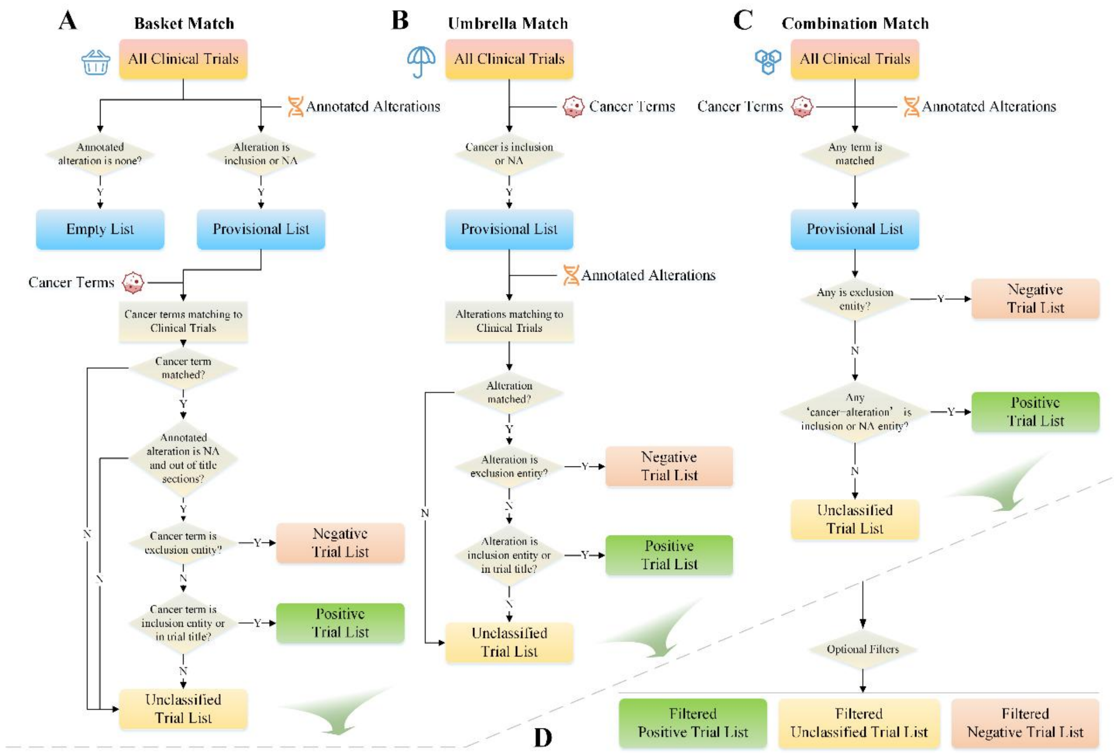


**Figure S2. Trials matching strategies.** A) Basket match prioritizes alterations as the primary matching condition; B) Umbrella match prioritizes cancer type as the primary matching condition; C) Combination match combines multiple matching conditions for more precise matching; D) Trial list filter allows users to filter and narrow down the list of matched clinical trials based on various criteria.
