## Supplementary File 3 for "OncoCTMiner: streamlining precision oncology trial matching via molecular profile analysis"

### Standard Processing Procedure for Tagging Clinical Trials

#### 1. Entity Categories

There are thirteen kinds of entities. Disease, gene, mutation, biomarker, chemical and therapy are manually tagged and reviewed, and the other entities are automatically annotated by the NLP-based tools.

E01. DISEASE (disease)

Focus on Cancer/Tumor/Neoplasm and other terminology for the same concepts.

E02. GENE (gene)

Gene and Protein.

E03.MUTATION (mutation)

Multi-omics alterations, which include:

| **Alter Type** | **Description** |
|---|---|
| OEXP | over expression |
| UEXP | under expression |
| NEXP | no expression |
| EXP | expression |
| RAM | rearrangement (or named fusion) |
| WT | wild-type |
| MSM | missense mutation |
| NSM | nonsense mutation |
| FS | frameshift mutation |
| DUP | duplication, including amplications |
| INDEL | single base to multiple bases, multiple bases to multiple bases, or multiple bases to single base |
| INS | insertion, either short or long insertion |
| DEL | deletion, either short or long deletion (including loss) |
| DMET | demethylation |
| HMET | hypermethylation |
| MET | methylation |
| AND | co-alterations |

| | |
|---|---|
| EGP | epigenetic phosphorylation |
| SNV | single reference base, and single variant base |
| CNV | copy number variation, not explicitly stating copy number deletion or copy number duplication |
| MUT | mutation, including alteration group and alteration that cannot be classified to any other type |
| OTH | alteration types that cannot be classified to any others and not suitable for mut |
| OR | either alteration |
| NA | unclassified alteration |

E11. BIOMARKER (biomarker)

NGS-related biomarker

*MSI:*

MSS (microsatellite stable)

MSI-L (microsatellite instability-low/MSI-low)

MSI-H (microsatellite instability-high/MSI-high)

*TMB:*

TMB-L (tumor mutational burden-low/low TMB/low tumor mutation burden/low tumor mutational burden)

TMB-M (tumor mutational burden-medium/medium TMB/medium tumor mutation burden/medium tumor mutational burden)

TMB-H (tumor mutational burden-high/high TMB/high tumor mutation burden/high tumor mutational burden)

IHC-related biomarkers

*MMR:*

dMMR (mismatch repair deficient/deficient mismatch repair)

pMMR (mismatch repair proficient/proficient mismatch repair)

E04. CHEMICAL (chemical)

Agency-approved drugs, including anti-cancer drugs and other types of drugs. Chemical substances or compounds under investigation.

E12. THERAPY (therapy)

All therapeutic methods in addition to the chemicals mentioned above.

E07. ADR (adr, reserved)

E08. SPECIES (species, reserved)

E09. ANATOMY (anatomy, reserved)

E10. RNA (rna, reserved)

E05. CLINSIG (clinsig, reserved)

E06. EVIDIRT (evidirt, reserved)

E13. PHENOTYPE (phenotype, reserved)

Including all diseases except cancer, and various symptoms or phenotypes.

#### 2. Eligibility Criteria Types

The critical requirements that people who want to participate in a clinical trial must meet or the characteristics they must have. Eligibility criteria consist of both inclusion criteria (which are required for a person to participate in the trial) and exclusion criteria (which prevent a person from participating). Types of eligibility criteria include whether a study accepts healthy volunteers, has age or age group requirements, or is limited by sex.

OncoCTMiner focus on the mining of precision oncology entities from all sections of each clinical trial from ClinicalTrials.gov (https://clinicaltrials.gov/). Each entity was tagged as one of the following criterias:

##### 2.1 Inclusion Criteria (inclusion)

A type of eligibility criteria. These are reasons that a person is allowed to participate in a clinical trial.

##### 2.2 Exclusion Criteria (exclusion)

A type of eligibility criteria. These are reasons that a person is not allowed to participate in a clinical trial.

##### 2.3 Not Criteria (not criteria)

Entities do not meet the inclusion or exclusion criteria mentioned in the section on eligibility criteria.

##### 2.4 NA (not available)

Criteria for entities cannot be categorized or do not need to be classified.

#### 3. Trial Tags

Trial tags are used to categorize clinical trial types. Since we are primarily interested in precision oncology, we must look for cancer therapies related clinical trials using biomarkers and mutations as eligibility criteria.

**Note:** For each tag, the related entity must appear in the text.

##### 3.1 Tag 1: Cancer

At least one of the condition of the clinical trial is ‘cancer/tumor/neoplasm’.

##### 3.2 Tag 2: Drug

The type of clinical trial is a drug intervention trial.

##### 3.3 Tag 3: Alteration

All NGS or IHC-related biomarkers and mutations can be classified as this type of tag.

##### 3.4 Tag 4: Gene

Gene or protein appear in the text.

The entity ‘gene’ is not tagged in the text, because ‘EGFR mutation’ is a mutation type of the EGFR gene, two tags of ‘Gene’ and ‘Alteration’ can be chosen simultaneously (Figure S1). Because of the appearance of ‘MSI’ belonging to biomarkers and ‘CRC’ belonging to cancers in the text, the tags of the clinical trial are ‘Cancer’ and ‘Alteration’ (Figure S2).

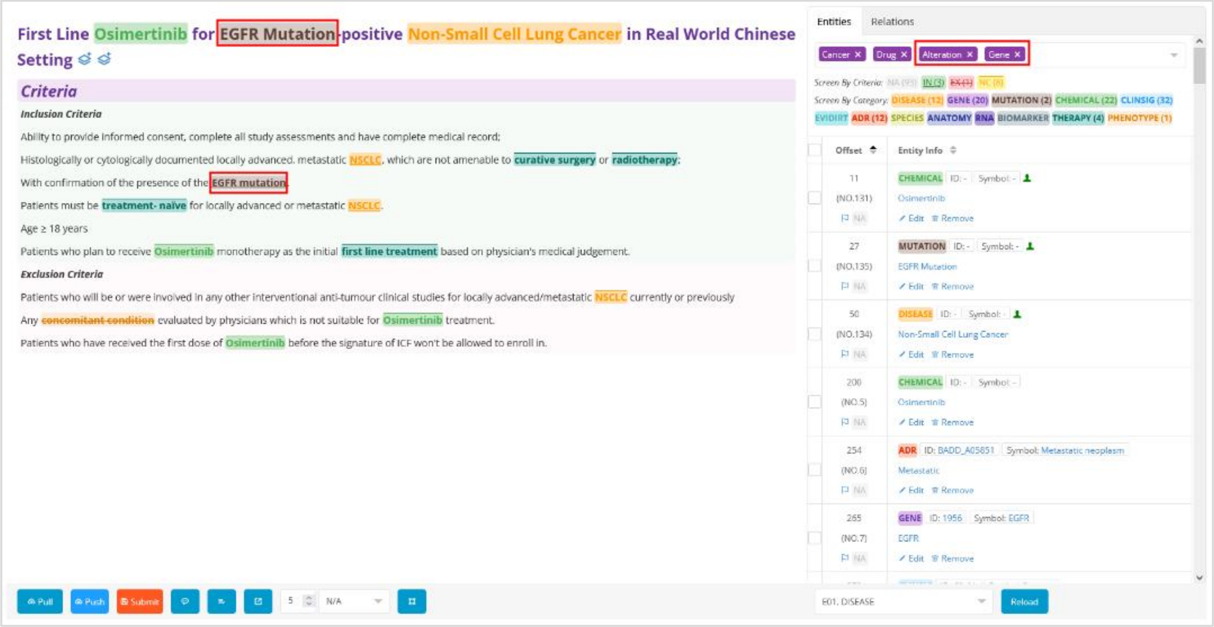

Figure S1. Trial tags choosing Alteration and Gene

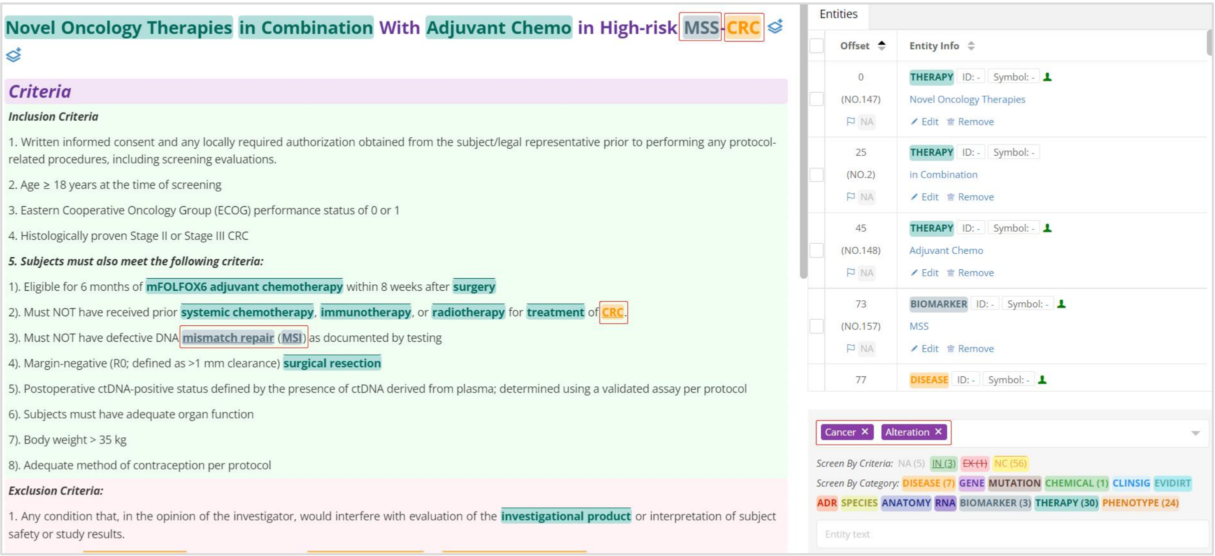

Figure S2. Trial tags choosing Cancer and Alteration

#### 4. Tagging rules

The following scenarios are only applicable to the current system version (v1).

##### 4.1 For text content

All sections from a clinical trial are needed for manual tagging.

##### 4.2 For entity categories

Only E01. DISEASE (disease), E02. GENE (gene), E03. MUTATION (mutation), E11. BIOMARKER (biomarker), E04. CHEMICAL (chemical), and E12. THERAPY (therapy) are needed for manual tagging.

##### 4.3 For criteria types

*Inclusion criteria*: An entity that meets the criteria is a prerequisite for inclusion in a clinical trial. To minimize missing eligible clinical trials, we used a 'loose access strategy' for the inclusion criteria mainly for 'disease', 'mutation' and 'biomarker' entities, which means that the entities are classed as inclusion as long as they fulfill the inclusion criterion, regardless of whether they have additional auxiliary conditions.

*Exclusion criteria*: An entity that meets the criteria is a prerequisite for exclusion from a clinical trial. To minimize missing eligible clinical trials, we used a 'rigorous exclusion strategy' for the exclusion criteria mainly for 'disease', 'mutation' and 'biomarker' entities, which means that the trial was not permitted to enroll as long as the entity was matched regardless of other circumstances. Only entities that match this requirement can be designated as exclusion criteria.

**Note:** If exclusion criteria contain any other preconditions, they cannot be grouped into exclusion categories, and only can be classified as not criteria.

*Not criteria*: Entities that do not meet the inclusion or exclusion criteria are just mentioned in the text. The entities which are belong to 'chemical', 'therapy' and 'gene' are grouped into not criteria categories in normal situations unless they can be grouped into inclusion category or exclusion category without any constraints.

*NA*: In general, only entities outside the 'Eligibility criteria' section are classified as NA, indicating the entity cannot be classified.

4.4 For standard of entity tagging

'Minimality' principle. The tagged entity is as short as possible on the base of expressing a complete thought after being standardized processed normally by the procedure. While the meaning of the entity which cannot be changed is the prerequisite (Figure S3).

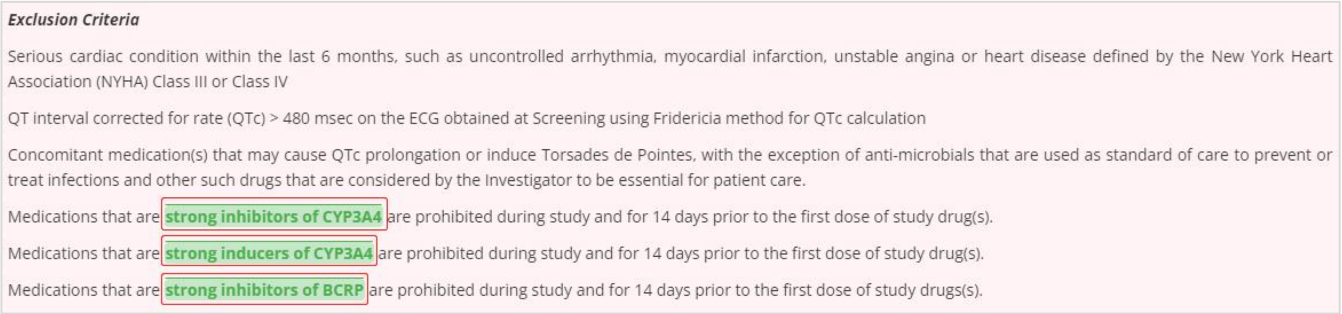

***Exclusion Criteria***

Serious cardiac condition within the last 6 months, such as uncontrolled arrhythmia, myocardial infarction, unstable angina or heart disease defined by the New York Heart Association (NYHA) Class III or Class IV

QT interval corrected for rate (QTc) > 480 msec on the ECG obtained at Screening using Fridericia method for QTc calculation

Concomitant medication(s) that may cause QTc prolongation or induce Torsades de Pointes, with the exception of anti-microbials that are used as standard of care to prevent or treat infections and other such drugs that are considered by the Investigator to be essential for patient care.

Medications that are strong inhibitors of CYP3A4 are prohibited during study and for 14 days prior to the first dose of study drug(s).

Medications that are strong inducers of CYP3A4 are prohibited during study and for 14 days prior to the first dose of study drug(s).

Medications that are strong inhibitors of BCRP are prohibited during study and for 14 days prior to the first dose of study drugs(s).

Figure S3. A kind of chemical (NCT04418167)

The entity is broken up into several entities when it contains multiple entities. For example, Two drugs in combination need to be broken up into two single drugs (Figure S4).

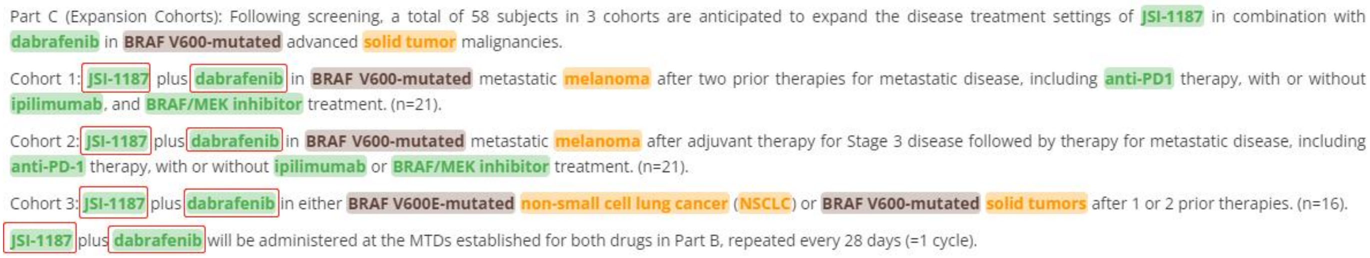

Part C (Expansion Cohorts): Following screening, a total of 58 subjects in 3 cohorts are anticipated to expand the disease treatment settings of JSI-1187 in combination with dabrafenib in BRAF V600-mutated advanced solid tumor malignancies.

Cohort 1: JSI-1187 plus dabrafenib in BRAF V600-mutated metastatic melanoma after two prior therapies for metastatic disease, including anti-PD1 therapy, with or without ipilimumab, and BRAF/MEK inhibitor treatment. (n=21).

Cohort 2: JSI-1187 plus dabrafenib in BRAF V600-mutated metastatic melanoma after adjuvant therapy for Stage 3 disease followed by therapy for metastatic disease, including anti-PD-1 therapy, with or without ipilimumab or BRAF/MEK inhibitor treatment. (n=21).

Cohort 3: JSI-1187 plus dabrafenib in either BRAF V600E-mutated non-small cell lung cancer (NSCLC) or BRAF V600-mutated solid tumors after 1 or 2 prior therapies. (n=16).

JSI-1187 plus dabrafenib will be administered at the MTDs established for both drugs in Part B, repeated every 28 days (=1 cycle).

Figure S4. Chemical ISI-1187 and chemical Dabrafenib combination (NCT04418167)

The entity belongs to a proper noun with no need for tagging (Figure S5). In general, the tagged entity subject must be a noun, which can't just be objective, verb or other non-noun (Figure S7). The entities outside the 'Eligibility criteria' section are grouped into NA category, for example, entities in the title are classified as NA (Figure S6).

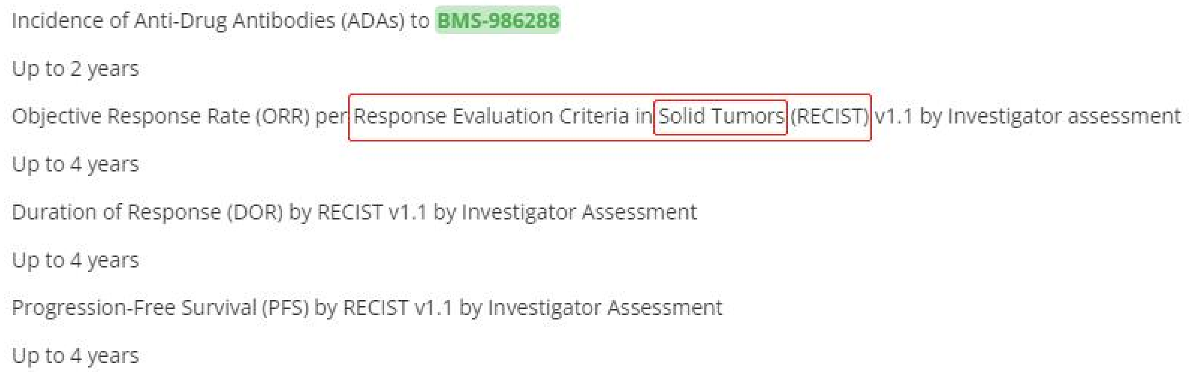

Figure S5. RSCIST a proper noun (NCT03994601)

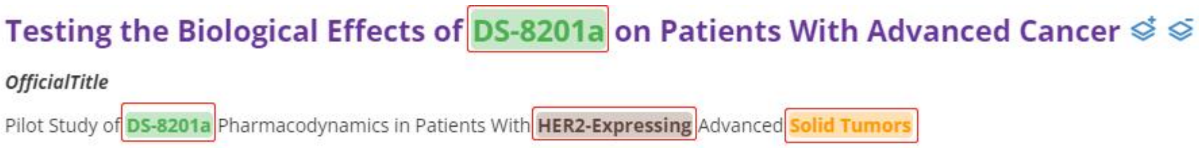

Figure S6. DS-8201a, HER2-Expressing and Solid Tumors grouped into NA (NCT04294628)

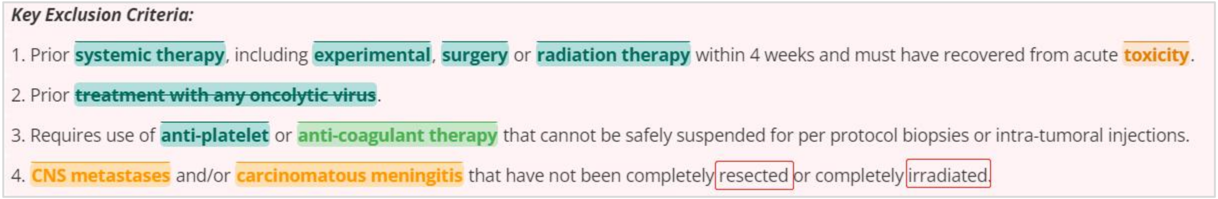

Figure S7. Verbs resected and irradiated (NCT04301011)

When an entity contains two parentheses, both of which are tagged or untagged (Figure S8). If the parenthesis is in the middle of two entities, the two entities must be tagged separately (Figure S9). About entity the order of preference is disease, mutation, biomarker and chemical over therapy over gene. If an entity belongs to a chemical, it will be tagged as chemical rather than therapy.

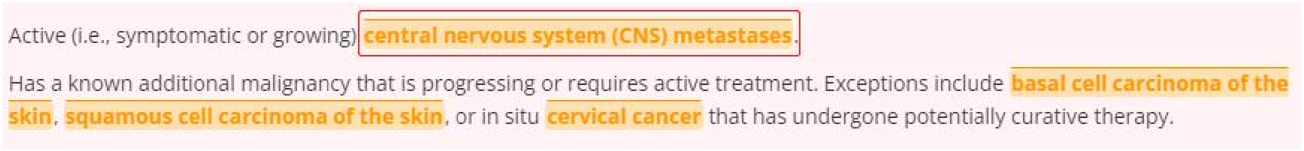

Figure S8. Parenthesis in the middle of the cancer (NCT04401995)

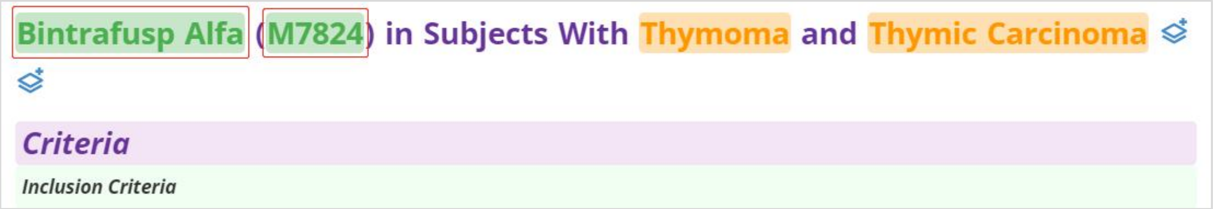

Figure S9. Parenthesis at the end of the chemical (NCT04417660)

4.5 For standard of disease tagging

4.5.1 For criteria types

In general, ‘Liver metastases’ is grouped into not criteria category unless there is supplementary information supporting it being grouped into inclusion category (Figure S10). ‘Liver involvement’ is similar to ‘Liver metastases’ (Figure S11). There is no need to tag tumor TNM stages as the disease (Figure S12).

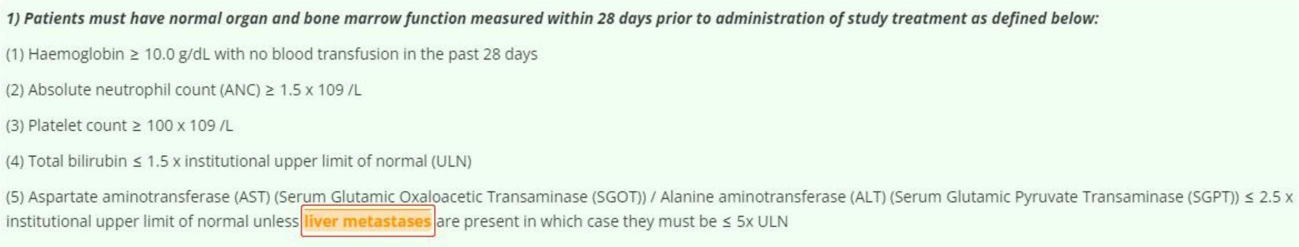

Figure S10. Liver metastases grouped into not criteria (NCT04150562)

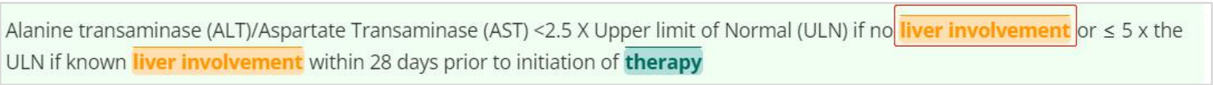

Figure S11. Liver involvement grouped into not criteria (NCT04692155)

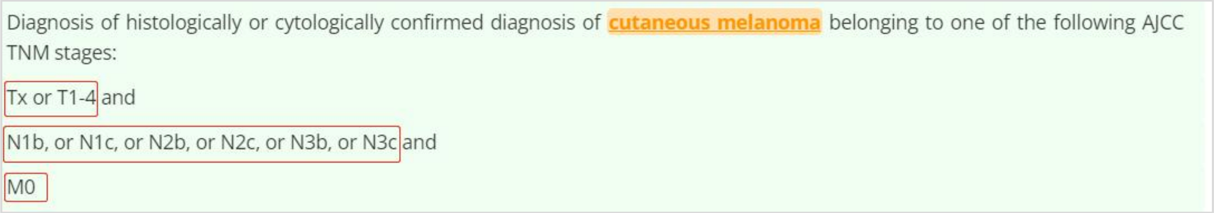

Figure S12. TNM stages not belonging to entity (NCT04401995)

##### 4.5.2 For disease tagging

Except for ‘acute’ and ‘chronic’, the following disease determiners do not need to be tagged: ‘clinically significant, clinically unsignificant, uncontrolled, active, inactive, advanced, refractory, high-risk, low-grade, high-grade, risk, archival, locally, symptomatic, asymptomatic, recurrent, in situ, primary, malignant, stage I/II/III/IV, early, mature, prior, primary, regional, aggressive, infiltrating, infiltrating, multicentric, indolent, bilateral’.

Words and phrases such as ‘metastasis, metastases, malignancy, lesion, primary, second primary, involvement’ appearing alone in the text should not be tagged as disease. In addition, ‘MRD(minimal residual disease)’ also should not be tagged as disease. The plus sign after the disease dose not need to be tagged (Figure S13). Words such as ‘metastatic, involvement, advanced, malignant, measurable’ with ‘disease’ or ‘lesion’ appearing at the same time should not be tagged as disease (Figure S14).

9). Patients must have a life expectancy ≥ 16 weeks.

10). At least one lesion, not previously irradiated, that can be accurately measured at baseline as ≥ 10 mm in the longest diameter (except lymph nodes which must have short axis ≥ 15 mm) with computed tomography (CT) or magnetic resonance imaging (MRI) and which is suitable for accurate repeated measurements.

Figure S13. Lesion not belonging to an entity (NCT04166435)

2. Adequate hematologic and organ function within 14 days before the first study treatment on Day 1 of Cycle 1.
3. Life expectancy of at least 6 months.
4. Measurable disease according to RECIST v1.1.
5. Eastern Cooperative Oncology Group (ECOG) Performance Status of 0 or 1.

Figure S14. Measurable disease not belonging to an entity (NCT04177108)

If the entities are composed of a disease, ‘in, for, of, from’, and a type of organ or cell, both the organ/cell and the disease should be tagged as a whole (Figure S15). If the entities are composed of a disease, ‘in, for, of, from’, and multiple organs or cells, both the organ/cell and the disease of the entities should be tagged as a whole (Figure S16). In addition, both the organ/cell and the disease of the entities that contain the disease, regardless of the number of types of organs and cells, should be tagged as a whole (Figure S17).

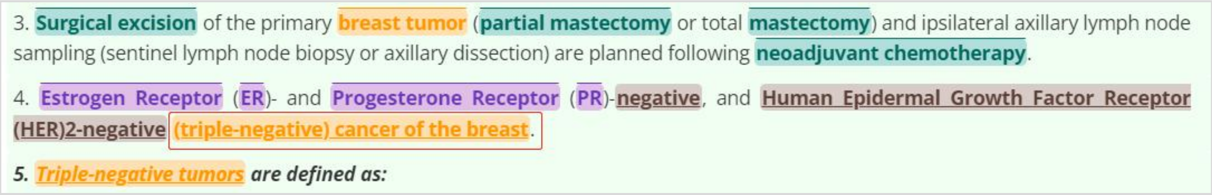

Figure S15. A kind of disease (NCT04083963)

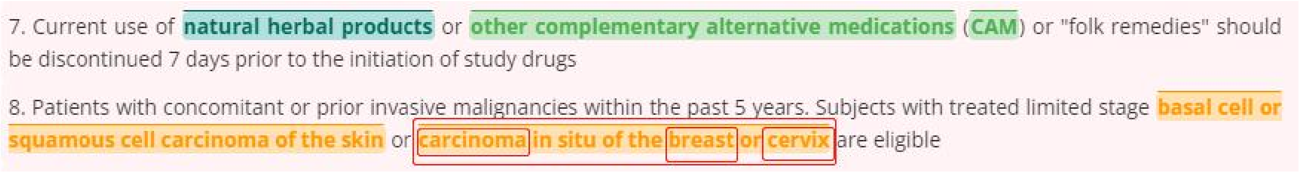

Figure S16. A kind of disease (NCT04090567)

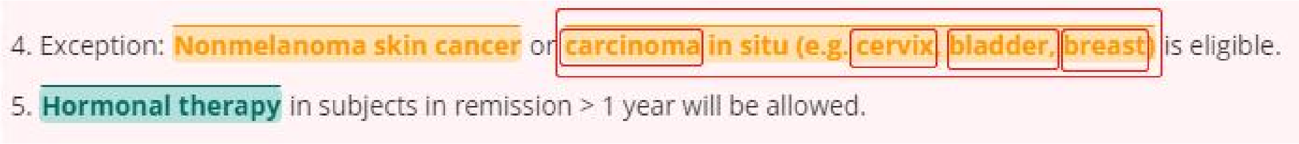

Figure S17. A kind of disease (NCT04088890)

4.6 For standard of chemical tagging

4.6.1 For criteria types

Inclusion criteria, there must be clear message that supports the chemical to be grouped into inclusion category (Figure S18). Exclusion criteria, the chemical should not be given before or during the clinical trial without any constraints (Figure S19). Not criteria, The chemical with any constraints including time, disease, dosage, whether combined with other chemicals, and any chemical determiners except for ‘strong, moderate, long-acting, short-acting’ should be grouped into not criteria category (Figure S20). The chemical may induce allergic reaction or subjects are unable/unwilling to take it or just intolerance, or the chemical which does not belong to inclusion category or exclusion category are grouped into not criteria category.

2. Patient must have progressed on, be intolerant of, decline, or be ineligible for, all available standard of care therapies

3. Part 2: locally advanced or metastatic NSCLC with KEAP1, NFE2L2 and/or STK11 mutation; Patients must have received at least a platinum doublet chemotherapy and an anti-PD-(L)1 antibody; Received up to 3 lines of systemic anticancer therapy in the recurrent or metastatic setting

Figure S18. ‘Must’ supporting inclusion category (NCT04471415)

*Exclusion Criteria*

1. Patient must not have received any prior systemic therapy for stage IV NSCLC. Patients must not have received prior anti-PD-1 or anti-PD-L1.

Figure S19. ‘Must not’ supporting exclusion category (NCT04470674)

9). In the investigator's judgment, the subject is unlikely to complete all protocol required study visits or procedures, including follow up visits, or comply with the study requirements for participation.

10). Primary immunodeficiency or history of autoimmune disease (e.g. Crohns, rheumatoid arthritis, systemic lupus) requiring systemic immunosuppression/systemic disease modifying agents within the last 2 years.

Figure S20. ‘Last 2 years’ supporting not criteria category (NCT04088890)

##### 4.6.2 For chemical tagging

The entity contains gene and chemical, such as ‘EGFR TKI/inhibitors/agonists/induces/targeting agent/targeting drugs/targeting therapy/directed therapy’ and so on, which is wholly tagged as chemical, it is the same case with entities containing anti-gene and therapy (Figure S21).

14. •Subjects with ALL: CD22 positive expression on malignant cells is required and must be detected by immunohistochemistry or flow cytometry. The choice of whether to use flow cytometry or immunohistochemistry will be determined by what is the most easily available tissue sample in each subject.

15. CD22 expression must be demonstrated subsequent to any anti-CD22 targeted therapy (e.g. Moxetumomab pasudotox or inotuzumab ogozamicin) in subjects with ALL.

Figure S21. Anti-CD22 targeted therapy belonging to chemical (NCT04088890)

The entity contains a kind of drug, of/for, and a single gene should be wholly tagged as chemical such as inhibitor of EGFR. If the entity contains a kind of drug, of/for, and multiple genes, each gene should be tagged as gene and drug should be tagged as chemical (Figure S22).

22. Patients may not receive concomitant chemotherapy, immunotherapy, or radiotherapy (other than as pertained to standard of care for GBM) while patients are on study

23. Prior treatment with ~~DNA damage response inhibitors~~ (including ~~inhibitors~~ of PARP, ATR, WEE)

Figure S22. Three kinds of chemical (NCT04555577)

The entity containing drug and therapy/treatment should be tagged as chemical as a whole. While therapy/treatment will be neglected during standardizing. The entity containing 'anticonvulsants/antibiotic/antihypertensive' and 'therapy' is tagged as chemical in on the whole. If the entity is end with 'agents/drugs/medications', it is also tagged as chemical (Figure S23). The chemical determiners such as 'second generation/first generation/strong/moderate/systemic' need to be tagged in an entity (Figure S24).

***Exclusion Criteria:***

1. Prior receipt of a selective FGFR inhibitor for any indication or reason.
2. Prior receipt of an anti-PD-1, anti-PD-L1, or anti-PD-L2 agent, or with an agent directed to another co-inhibitory T-cell receptor.
3. Receipt of anticancer medications or investigational drugs for unresectable and/or metastatic disease.

Figure S23. The entity ending with agents (NCT04003610)

4. Patients with a condition requiring systemic treatment with either corticosteroids (> 10 mg daily prednisone or equivalent) or other immunosuppressive medications within 14 days of study drug administration. Inhaled or topical steroids are permitted in the absence of active autoimmune disease.

5. Prior therapy for melanoma with the following exceptions which are allowed: 1) surgery for the melanoma lesion(s), 2) adjuvant RT after neurosurgical resection for CNS lesions or for resected locoregional disease, and 3) prior adjuvant IFN-alpha, ipilimumab and nivolumab (see qualifier below).

Figure S24.The entity containing systemic (NCT03999749)

Words and phrases such as 'oral/injection/infusion/intravenous injection(iv)/inhaled' appearing conjunction with chemical do not need to be tagged into the entity (Figure S25). 'Hormone/estrogen/corticosteroid replacement therapy' should be tagged as chemical in whole (Figure S26), and the entity only contains the drug of chemotherapy/radiotherapy is the same as above. 'Tablets/biological/medical/equivalent/medication' don't need to be tagged when it appears alone. The phrase of hypersensitivity to similar chemical also don't need to be tagged.

5. Any illness or medical condition that is unstable or could jeopardize the safety of the subject or her compliance with study requirements.

6. Subjects unable to swallow oral medications.

Figure S25. Oral medications not belonging to chemical (NCT04551495)

7. a. Replacement therapy (e.g. thyroxine, insulin, or physiologic corticosteroid replacement therapy for adrenal or pituitary insufficiency) is not considered a form of systemic treatment and is allowed.

8. Patients who are receiving daily systemic corticosteroids that are above physiological doses for any reason or who are under immunosuppressive or immunomodulatory treatment

Figure S26. Physiologic corticosteroid replacement therapy belonging to chemical (NCT04577326)

##### 4.6.3 White list

Platinum, insulin, thyroxine.

##### 4.6.4 Black list

Co (Figure S27), bilirubin, creatinine (Figure S28), alanine, aspartate, placebo.

**Evaluation of Co-formulated Pembrolizumab/Quavonlimab (MK-1308A) Versus Other Treatments in Participants With Microsatellite Instability-High (MSI-H) or Mismatch Repair Deficient (dMMR) Stage IV Colorectal Cancer (CRC) (MK-1308A-008)**

***Criteria***

Figure S27. Prefix co not belonging to entity (NCT04895722)

***27. Adequate renal, hepatic, pulmonary and cardiac function defined as:***

1). Creatinine within institutional norms for age (i.e. ≤ 2 mg/dL in adults or according to table below in children <18 years) OR creatinine clearance (as estimated by Cockcroft Gault Equation) ≥ 60 mL/min Age (Years) Maximum Serum Creatinine (mg/dL)

28. Years - Serum Creatinine (mg/dL) = 0.8; 5 < age ≤ 10 Years - Serum Creatinine (mg/dL) = 1.0; >10-18 years - Serum Creatinine (mg/dL) = 1.2; > 18 years - Serum Creatinine (mg/dL) = 2.0

29. Serum alanine aminotransferase/aspartate aminotransferase(ALT/AST)≤ 10x upper limit of normal (ULN) (unless elevated ALT/AST is associated with leukemia involvement of the liver, in which case this criterion will be waived and not disqualify a patient).

30. Total bilirubin ≤ 1.5 mg/dl, except in subjects with Gilbert's syndrome.

Figure S28. Creatinine not belonging to entity (NCT04088864)

4.7 For standard of therapy tagging

4.7.1 For criteria types

Inclusion criteria, there must be clear message that supports the therapy to be grouped into inclusion category (Figure S29). Exclusion criteria, the therapy should not be given before or during the clinical trial without any constraints.

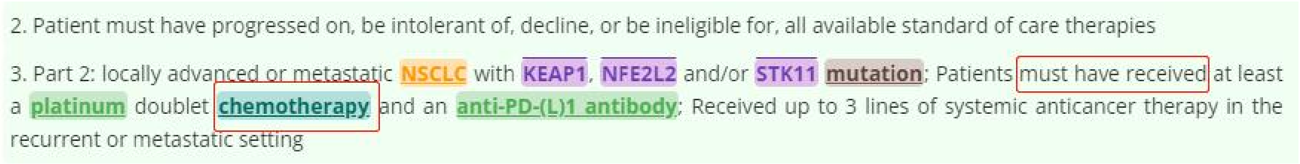
2. Patient must have progressed on, be intolerant of, decline, or be ineligible for, all available standard of care therapies

3. Part 2: locally advanced or metastatic NSCLC with KEAP1, NFE2L2 and/or STK11 mutation; Patients must have received at least a platinum doublet chemotherapy and an anti-PD-(L)1 antibody; Received up to 3 lines of systemic anticancer therapy in the recurrent or metastatic setting

Figure S29. ‘Must’ supporting inclusion category (NCT04471415)

4.7.2 For therapy tagging

‘antiviral/antiretroviral’ should be tagged as therapy, so are the same case of ‘resection, transplantation, vaccines/vaccination, R-CHOP’ should be tagged as therapy (Figure S30). ‘Oral/injection/infusion/intravenous injection(iv)/inhaled/biopsy’ don't need to be tagged when it appears alone(Figure S31). Therapy methods with treatment/therapy should be wholly tagged as therapy apart from ‘(anti-)cancer treatment/therapy, definitive therapy, immunosuppressive therapy, curative therapy, neoadjuvant therapy’(Figure S32).

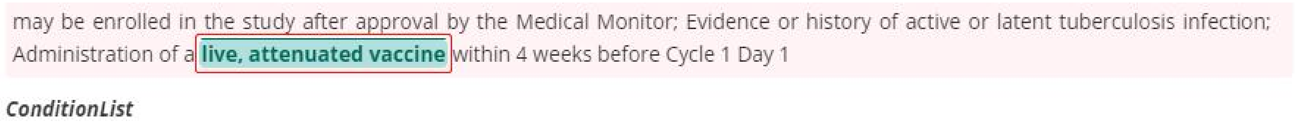
may be enrolled in the study after approval by the Medical Monitor; Evidence or history of active or latent tuberculosis infection; Administration of a live, attenuated vaccine within 4 weeks before Cycle 1 Day 1

*ConditionList*

Figure S30. A kind of therapy (NCT04189614)

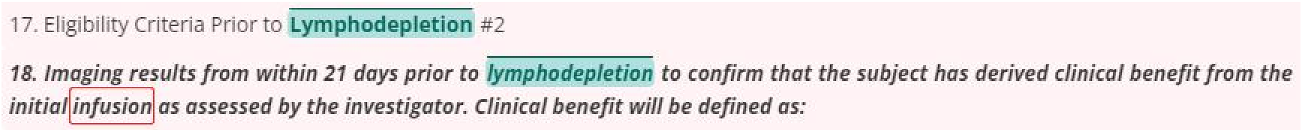
17. Eligibility Criteria Prior to Lymphodepletion #2

*18. Imaging results from within 21 days prior to lymphodepletion to confirm that the subject has derived clinical benefit from the initial infusion as assessed by the investigator. Clinical benefit will be defined as:*

Figure S31. Infusion not belonging to therapy (NCT04083495)

For participants treated with neoadjuvant therapy and HER2-targeted therapy: A minimum of 4 cycles of T-DM1 in the adjuvant setting. NOTE: Participants may have received up to approximately 6 cycles of adjuvant trastuzumab prior to initiation of T-DM1. Additionally, participants may have switched to trastuzumab-based therapy (monotherapy or in combination with other HER2-targeted therapies) after 4 cycles of T-DM1

Figure S32. Neoadjuvant therapy not belonging to therapy (NCT04752332)

‘Chemotherapy’ should be tagged as therapy, while specific chemotherapeutic drug should be tagged as chemical (Figure S33). In general, XXX-based chemotherapy should be tagged as therapy. The term ‘regimen’ used alone or with other determiners does not need to be tagged. Specific therapy and chemical should be tagged separately, for example, the first part ‘XXX’ of ‘XXX-based chemotherapy’ should be tagged as chemical, while the last part should be tagged as therapy; Organs don't need to be tagged in sentences containing multiple organs and therapy. Fruit words do not need to be tagged.

Patients with EGFR mutations (deletions in exon 19 and L858R in exon 21 of the EGFR gene), plan to receive First-generation EGFR-TKIs (Gefitinib, Erlotinib, Icotinib) or third generation EGFR-TKIs (including but not limited to Osimertinib, Almonertinib, Furmonertinib) monotherapy for the first time (patients who have been using first- or third-generation EGFR-TKIs for less than 28 days can be enrolled).

Patients positive for EGFR gene mutation (deletions in exon 19 and L858R in exon 21 of the EGFR gene), with disease progression after receiving chemotherapy can be enrolled.

Figure S33. Chemotherapy belonging to therapy (NCT04401059)

4.7.3 White list

CAR-T (Figure S34), R-CHOP, chemotherapy.

12). Subject has not received anti-CD30 antibody-based therapy within the previous 4 weeks prior to lymphodepletion.

13). Subject has not received chemotherapy within the previous 3 weeks prior to lymphodepletion.

14). Subject does not have rapidly progressive disease, per treating oncologist's discretion.

15). Subject is a good candidate for CAR T cell therapy, per treating oncologist's discretion.

Figure S34. ‘CAR T’ belonging to therapy (NCT04083495)

4.7.4 Black list

immunohistochemical/immunohistochemistry.

##### 4.8 For standard of mutation tagging

###### 4.8.1 For criteria types

Tagging rules has described how mutations are grouped.

###### 4.8.2 For mutation tagging

According to ‘minimality’ principle, if the entity contains mutation and disease, each part of the entity need to be tagged separately (Figure S35). If the entity contains a gene and ‘mutation(s)/express(ion)/aberration(s)/translocation’, it need to be tagged as mutation in whole (Figure S36). If the entity contains multiple genes and ‘mutation(s)/express(ion)/aberration(s)/translocation’, each part of the entity need to be tagged separately, that is gene is tagged as gene, mutation is tagged as mutation (Figure S37).

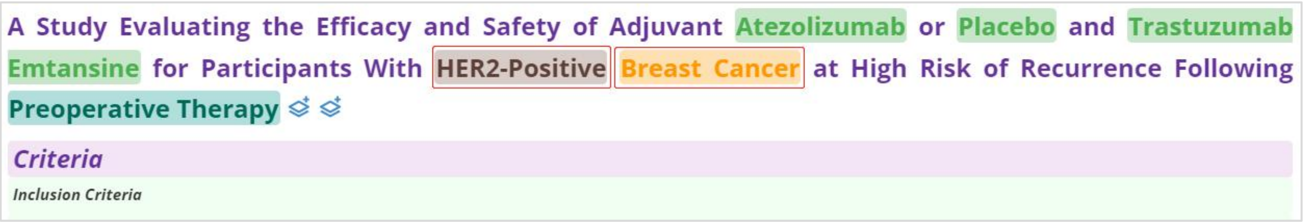

Figure S35. Two kinds of entities (NCT04873362)

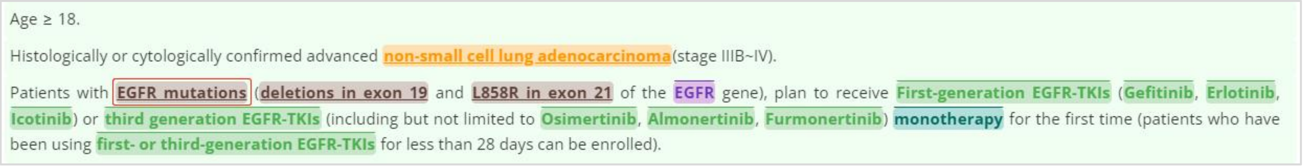

Figure S36. A kind of mutation (NCT04401059)

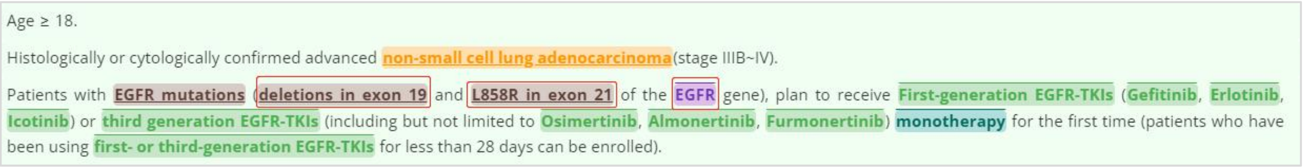

Figure S37. Two kinds of mutation (NCT04401059)

If the entity just contains gene and 'positive/negative' without mutation in the back, it should be tagged as mutation in whole (Figure S38). If there is a plus at the back of gene, both of which should be tagged as mutation in whole (Figure S39). HRD, sytogenetic abnormality should be tagged as mutation, in addition, mutation in exon X also should be tagged as mutation.

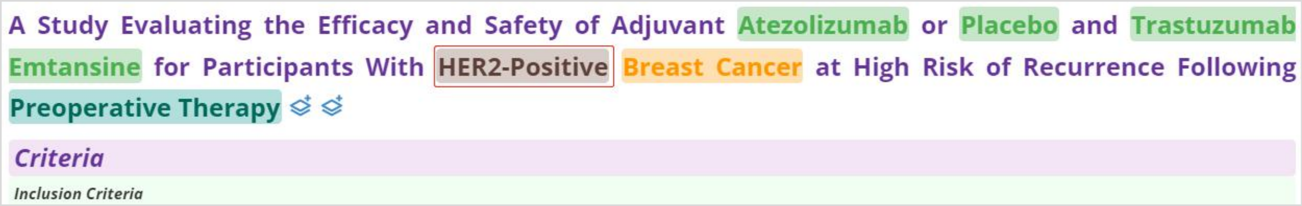

Figure S38. A kind of mutation (NCT04873362)

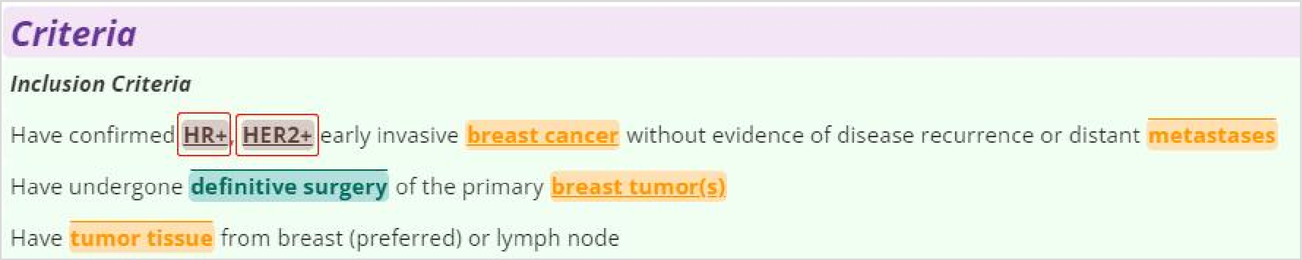

Figure S39. HR+/HER2+ belonging to mutation (NCT04752332)

4.9 For standard of biomarker tagging

4.9.1 For criteria types

Criteria types of biomarker are the same as mutation.

4.9.2 For biomarker tagging

All biomarker entities are tagged as described in part E11 BIOMARKER.

##### 4.10 For standard of gene tagging

###### 4.10.1 For criteria types

In general, genes are grouped into not criteria category unless there is clear message for inclusion category or exclusion category (Figure S40).

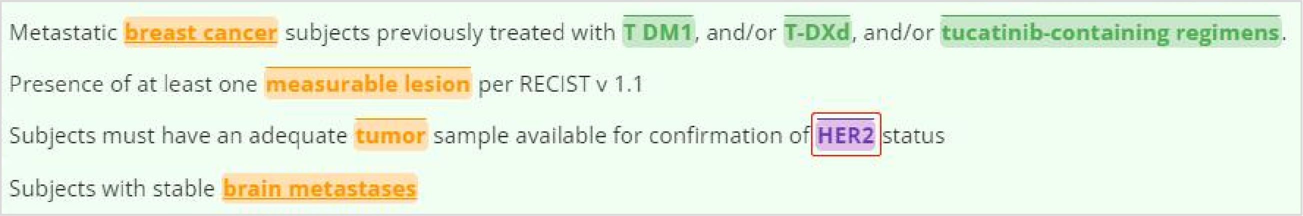
Metastatic breast cancer subjects previously treated with T DM1, and/or T-DXd, and/or tucatinib-containing regimens.

Presence of at least one measurable lesion per RECIST v 1.1

Subjects must have an adequate tumor sample available for confirmation of HER2 status

Subjects with stable brain metastases

Figure S40. HER2 grouped into not criteria (NCT04829604)

###### 4.10.2 Black list

FISH, MRI, CAP, AST, ALT, min/1, GFR, PET/CT, AEs, PCR, ANC, eGFR, ARM, ICF, DTR, CSF, ISH (FigureS41).

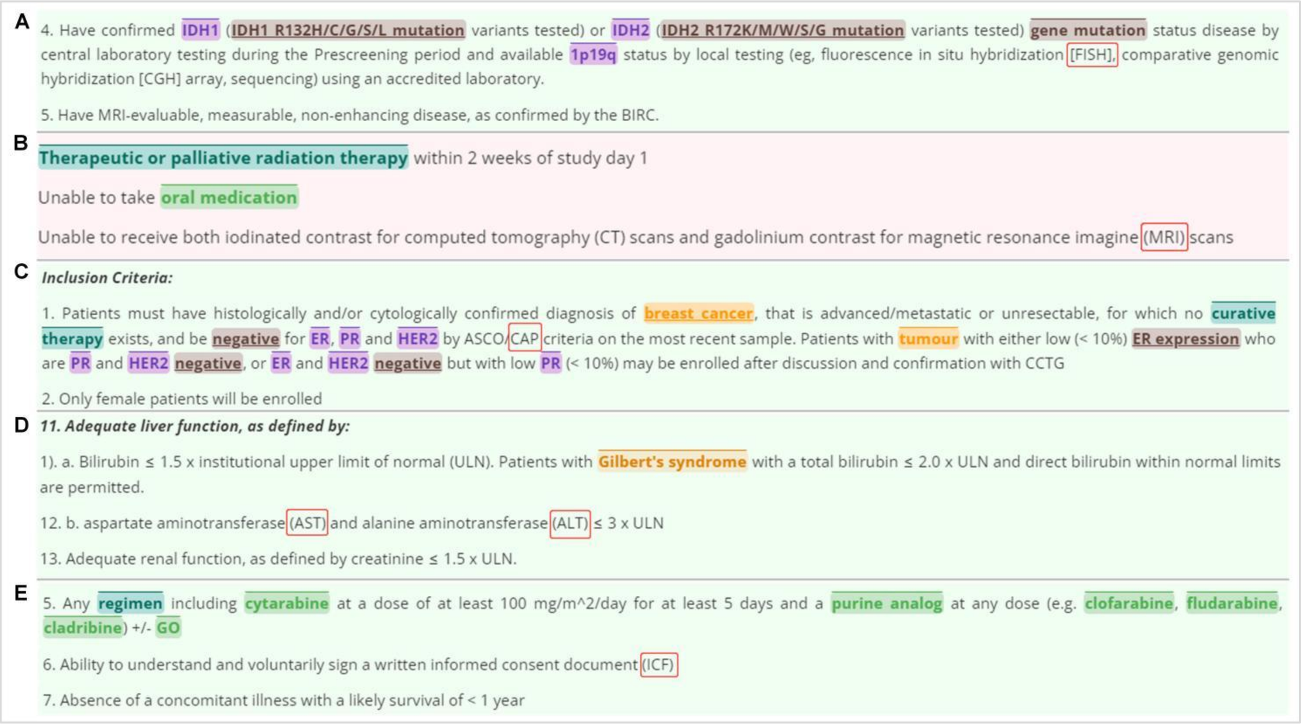
A

4. Have confirmed IDH1 (IDH1 R132H/C/G/S/L mutation variants tested) or IDH2 (IDH2 R172K/M/W/S/G mutation variants tested) gene mutation status disease by central laboratory testing during the Prescreening period and available 1p19q status by local testing (eg, fluorescence in situ hybridization [FISH], comparative genomic hybridization [CGH] array, sequencing) using an accredited laboratory.

5. Have MRI-evaluable, measurable, non-enhancing disease, as confirmed by the BIRC.

B

Therapeutic or palliative radiation therapy within 2 weeks of study day 1

Unable to take oral medication

Unable to receive both iodinated contrast for computed tomography (CT) scans and gadolinium contrast for magnetic resonance imagine (MRI) scans

C

*Inclusion Criteria:*

1. Patients must have histologically and/or cytologically confirmed diagnosis of breast cancer, that is advanced/metastatic or unresectable, for which no curative therapy exists, and be negative for ER, PR and HER2 by ASCO/CAP criteria on the most recent sample. Patients with tumour with either low (< 10%) ER expression who are PR and HER2 negative, or ER and HER2 negative but with low PR (< 10%) may be enrolled after discussion and confirmation with CCTG

2. Only female patients will be enrolled

D

***11. Adequate liver function, as defined by:***

1). a. Bilirubin ≤ 1.5 x institutional upper limit of normal (ULN). Patients with Gilbert's syndrome with a total bilirubin ≤ 2.0 x ULN and direct bilirubin within normal limits are permitted.

12. b. aspartate aminotransferase (AST) and alanine aminotransferase (ALT) ≤ 3 x ULN

13. Adequate renal function, as defined by creatinine ≤ 1.5 x ULN.

E

5. Any regimen including cytarabine at a dose of at least 100 mg/m^2/day for at least 5 days and a purine analog at any dose (e.g. clofarabine, fludarabine, cladribine) +/- GO

6. Ability to understand and voluntarily sign a written informed consent document (ICF)

7. Absence of a concomitant illness with a likely survival of < 1 year

Figure S41. FISH, MRI, CAP, AST, ALT, ICF not belonging to gene

*Last updated on 10-Jul-2023 15:47 (UTC+8)*
